# Comparative performance of five commercially available serologic assays to detect antibodies to SARS-CoV-2 and identify individuals with high neutralizing titers

**DOI:** 10.1101/2020.08.31.20184788

**Authors:** Eshan U. Patel, Evan M. Bloch, William Clarke, Yu-Hsiang Hsieh, Denali Boon, Yolanda Eby, Reinaldo E. Fernandez, Owen R. Baker, Morgan Keruly, Charles S. Kirby, Ethan Klock, Kirsten Littlefield, Jernelle Miller, Haley A. Schmidt, Philip Sullivan, Estelle Piwowar-Manning, Ruchee Shrestha, Andrew D. Redd, Richard E. Rothman, David Sullivan, Shmuel Shoham, Arturo Casadevall, Thomas C. Quinn, Andrew Pekosz, Aaron A.R. Tobian, Oliver Laeyendecker

## Abstract

Accurate serological assays to detect antibodies to SARS-CoV-2 are needed to characterize the epidemiology of SARS-CoV-2 infection and identify potential candidates for COVID-19 convalescent plasma (CCP) donation. This study compared the performance of commercial enzyme immunoassays (EIAs) to detect IgG or total antibodies to SARS-CoV-2 and neutralizing antibodies (nAb). The diagnostic accuracy of five commercially available EIAs (Abbott, Euroimmun, EDI, ImmunoDiagnostics, and Roche) to detect IgG or total antibodies to SARS-CoV-2 was evaluated from cross-sectional samples of potential CCP donors that had prior molecular confirmation of SARS-CoV-2 infection for sensitivity (n=214) and pre-pandemic emergency department patients for specificity (n=1,102). Of the 214 potential CCP donors, all were sampled >14 days since symptom onset and only a minority had been hospitalized due to COVID-19 (n=16 [7.5%]); 140 potential CCP donors were tested by all five EIAs and a microneutralization assay. When performed according to the manufacturers’ protocol to detect IgG or total antibodies to SARS-CoV-2, the sensitivity of each EIA ranged from 76.4% to 93.9%, and the specificity of each EIA ranged from 87.0% to 99.6%. Using a nAb titer cutoff of ≥160 as the reference positive test (n=140 CCP donors), the ***empirical*** area under receiver operating curve of each EIA ranged from 0.66 (Roche) to 0.90 (Euroimmun). Commercial EIAs with high diagnostic accuracy to detect SARS-CoV-2 antibodies did not necessarily have high diagnostic accuracy to detect high nAbs. Some but not all commercial EIAs may be useful in the identification of individuals with high nAbs in convalescent individuals.

## INTRODUCTION

Globally, as of August 2020, there have been over 23.5 million reported cases of Severe Acute Respiratory Syndrome Associated Coronavirus 2 (SARS-CoV-2) infection, which causes Coronavirus-19 (COVID-19) disease.^1^ Surveillance based on case-reporting is informative but it significantly underestimates the true burden of infection and can lead to biased epidemiological inferences. Accurate and reliable serological assays to detect SARS-CoV-2 antibodies can be used to better understand the epidemiology of SARS-CoV-2 infection at the population-level, as the presence of antibodies to SARS-CoV-2 indicates recent or prior exposure to the virus.^2^ Serological assays can also be useful for screening blood donations, qualifying individuals for convalescent plasma donation, monitoring immune responses to vaccine candidates, clinically managing patients, and studying the natural history of infection.^2^ It is still unknown whether the presence of antibodies against SARS-CoV-2 confers immunity against reinfection with the virus, or how long those antibodies persist following infection.

As of August 2020, 39 commercially available serological assays have received an individual emergency use authorization (EUA) by the US Food and Drug Administration (FDA) for the detection of antibodies to SARS-CoV-2.^3^ These assays detect IgA, IgM, IgG or total antibodies to the subunit 1 of the spike glycoprotein (S1), the spike glycoprotein receptor binding domain (RBD), or the recombinant nucleocapsid protein (N) of the virus. The assays can also be categorized, broadly, as (1) lateral flow immunoassays (LFAs); (2) enzyme-linked immunosorbent assays (ELISAs); and (3) chemiluminescent immunoassays (CLIAs). ELISAs and CLIAs (collectively known as enzyme immunoassays [EIAs]) often provide semiquantitative output that can be interpreted as antibody titers, whereas current LFAs are strictly qualitative. Recent systematic reviews of the literature have noted the need for additional data on the performance of commercially available SARS-CoV-2 serologic assays, as most previous studies have been deemed to have a high risk of bias, particularly due to the use of small sample sizes and/or exclusion of specimens from asymptomatic SARS-CoV-2 infections and mild or moderate cases of COVID-19.^4-6^

Commercial SARS-CoV-2 EIAs may have an additional role in the implementation of COVID-19 convalescent plasma (CCP) therapy programs.^2,7^ The FDA recently issued an EUA for CCP therapy.^8^ Indeed, observational evidence suggests CCP is likely safe and efficacious, particularly when administered early in the disease process.^9,12^ Higher IgG antibody titers to the S1 protein in CCP transfused to COVID-19 patients have been associated with decreased mortality.^12^ Higher IgG antibody titers to the spike (S) and nucleocapsid (N) protein of SARS-CoV-2 have also been shown to correlate with SARS-CoV-2 neutralizing antibody (nAb) titers^13-16^, which are presumed to be critical for viral clearance. Current in vitro assays to detect nAbs are resource- and time-intensive, and are not typically conducted in clinical laboratories. Commercial SARS-CoV-2 EIAs that are already in use to qualify CCP donors could also potentially be applied to identify those with high nAbs. However, data on the comparative performance of commercial SARS-CoV-2 EIAs to discriminate between CCP donors with high and low nAbs are limited.

This study was designed to compare the performance of five commercially available EIAs to detect IgG or total antibodies to SARS-CoV-2 and to discriminate between high and low nAbs.

## MATERIALS AND METHODS

### Ethics statement

This study used stored samples and data from two parent studies that were approved by The Johns Hopkins University School of Medicine Institutional Review Board. All samples were de-identified prior to laboratory testing. Both studies were conducted according to the ethical standards of the Helsinki Declaration of the World Medical Association.

### Study specimens

To test the clinical sensitivity of SARS-CoV-2 EIAs, we included stored plasma specimens from a convenience sample of potential CCP donors that were recruited in the Baltimore, MD and Washington DC area (n=214).^13^ Individuals were eligible for enrollment if they had a documented history of a positive molecular assay test result for SARS-CoV-2 infection (confirmed by medical chart review or shared clinical documentation) and met standard self-reported eligibility criteria for blood donation. Demographic information of included CCP donors is shown in ***Supplemental Table 1***. Among included CCP donors, there was a median of 44 days from diagnosis until sample collection (interquartile range, 38-50 days). Although all included CCP donors were symptomatic at the time of SARS-CoV-2 infection, less than 10% had a history of hospitalization due to COVID-19. To test the clinical specificity of SARS-CoV-2 EIAs, we included stored serum specimens from an identity-unlinked serosurvey conducted in 2016 among adult patients attending the Johns Hopkins Hospital Emergency Department (n=1,102). Both parent studies were cross-sectional and no individual contributed multiple specimens. All plasma/serum samples were stored at -80°C until assays were performed.

### SARS-CoV-2 EIAs

Plasma/serum specimens were analyzed using five commercially available EIAs: the Euroimmun Anti-SARS-CoV-2 ELISA, the Epitope Diagnostics, Inc. (EDI) Novel Coronavirus COVID-19 IgG ELISA Kit, the ImmunoDiagnostics SARS-CoV-2 NP IgG ELISA kit, the Abbott-Architect SARS-CoV-2 IgG chemiluminescent microparticle immunoassay (CMIA) and the Roche Diagnostics Elecsys®Anti-SARS-CoV-2 E-CLIA (**Table 1**). These commercially available EIAs were selected either because data on performance characteristics for the assay are limited and/or the assay has received an EUA by the FDA. The target antigen for each EIA is the nucleocapsid protein with the exception of the Euroimmun ELISA for which the target antigen is the S1 protein. The Roche assay measures total antibodies to SARS-CoV-2, whereas the others measure only IgG to SARS-CoV-2. EIAs were conducted according to the manufacturers’ instructions. The intended use of each EIA is the qualitative detection of antibodies; however, each EIA provides semi-quantitative output normalized by a calibrator. For simplicity, we refer to the normalized continuous output of each EIA as a “ratio” value. The manufacturers’ ratio cutoffs to qualitatively indicate seropositivity, indeterminate serostatus, or seronegativity for SARS-CoV-2 antibodies are provided (**Table 1**). Specimens were tested by each EIA based on sample volume availability and assay kit availability at the time of testing.

**Table 1.** Characteristics of commercial SARS-CoV-2 enzyme immunoassays evaluated.

| Manufacturer | Assay Name | Target antigen (recombinant) | Platform | Manufacturer's interpretation | No. Samples Evaluated |
| --- | --- | --- | --- | --- | --- |
| Euroimmun, Lubeck, Germany | Anti-SARS-CoV-2 ELISA (IgG) <sup>a</sup> | Spike-1 protein | Manual ELISA | Negative: S/C ratio < 0.8<br>Borderline: S/C ratio ≥0.8 & <1.1<br>Positive: S/C ratio ≥1.1 | CCP donors: 146<br>Pre-pandemic: 562 |
| Epitope Diagnostics, Inc., San Diego, CA | EDI™ Novel Coronavirus COVID-19 IgG ELISA Kit | Nucleocapsid protein | Manual ELISA | Negative: OD-n < 0.18<br>Borderline: OD-n ≥0.18 & <0.22<br>Positive: OD-n ≥0.22 | CCP donors: 146<br>Pre-pandemic: 579 |
| ImmunoDiagnostics Limited, Sha Tin, Hong Kong <sup>b</sup> | SARS-CoV-2 NP IgG ELISA kit | Nucleocapsid protein | Manual ELISA | Negative: OD-n < 0.15<br>Borderline: OD-n ≥0.25 & ≤0.50<br>Positive: OD-n > 0.50 | CCP donors: 140<br>Pre-pandemic: 306 |
| Abbott Laboratories Inc., Abbott Park, IL | Abbott-Architect SARS-CoV-2 IgG assay <sup>a</sup> | Nucleocapsid protein | Abbott Architect™ i2000 (CMIA) <sup>c</sup> | Negative: index (S/C) <1.40<br>Positive: index (S/C) ≥1.40 | CCP donors: 146<br>Pre-pandemic: 500 |
| Roche Diagnostics | Elecsys® Anti-SARS-CoV-2 <sup>a</sup> | Nucleocapsid protein | Roche cobas™ c 422 analyzer (ECLIA) | Non-reactive: index <1.0<br>Reactive: index ≥1.0 | CCP donors: 214<br>Pre-pandemic: 500 |

<sup>a</sup> This assay had received emergency use authorization by the US Food and Drug Administration prior to August 20, 2020.
<sup>b</sup> ImmunoDiagnostics recommends each lab create its own cut-offs for qualitative interpretation.
<sup>c</sup> This study utilized the Abbott Architect i1000sr platform.

### Microneutralization assay

Plasma nAb titers were quantified against 100 fifty percent tissue culture infectious doses (TCID50) using a microneutralization (NT) assay in VeroE6-TMPRSS2 cells, which has been previously described.^13,17^ In brief, plasma was diluted 1:20 and subsequent two-fold dilutions. Infectious virus was added to the plasma dilutions at a final concentration of 1×10^4^ TCID_50_/ml. After a 1-hour incubation at room temperature, 100μL of each sample dilution was added to 6 wells in a 96-well plate of VeroE6-TMPRSS2 cells,^18^ and incubated for 6 hours at 37°C. The inocula were removed from the plate, fresh media was added, and the plate was incubated at 37°C for 48 hours. The cells were fixed with 4% formaldehyde (in each well), incubated for 4 hours at room temperature, and stained with Napthol Blue Black (Sigma-Aldrich). We calculated a nAb titer area under the curve (AUC) value for each sample using the exact number of wells protected from infection at every dilution. Samples with no neutralizing activity were assigned a value of one-half the lowest measured AUC.

### Statistical analysis

The diagnostic accuracy of each EIA to detect IgG or total antibodies to SARS-CoV-2 was examined using CCP donor specimens as reference standard positive and pre-pandemic specimens as reference standard negative. For each EIA, non-parametric, empirical receiver operating curve analysis (ROC) was performed to calculate the area under the receiver operating curve (AUROC). This analysis was also done using the manufacturers’ cut-offs. Sensitivity (%) was calculated as 100 x (Positive/[Positive + False-Negative]). Specificity (%) was calculated as 100 x (Negative/[Negative + False-Positive]). For these analyses, an available-case approach was used for each EIA and indeterminate results were considered to be seronegative. Three separate sensitivity analyses were conducted: (1) we performed head-to-head comparisons, (2) we considered indeterminate specimens as positive, and (3) we excluded indeterminate specimens. Exact binomial (Clopper-Pearson) 95% confidence intervals (CI) were calculated for estimates.

The remaining analyses were conducted in CCP donors that had data for all five EIAs and nAb titers (n=140). The correlation of EIA ratios and nAb AUC values were examined using spearman’s correlation coefficients (ρ) with 95% CIs estimated over 1000 bootstrap iterations. We evaluated four binary cut-offs of the nAb AUC value to indicate “high” nAbs titers: ≥20, ≥40, ≥80, and ≥160. For each nAb AUC cut-off, we evaluated the performance of each EIA to discriminate between low and high nAb titers using empirical ROC analysis.

According to the recent EUA for CCP therapy, all CCP donors will be required to be antibody positive for SARS-CoV-2. Thus, we also calculated the positive percentage agreement and negative percentage agreement between each binary nAb threshold and each EIA using the manufacturer’s cut-offs originally recommended for SARS-CoV-2 serostatus in the CCP donor population (indeterminates were considered as seronegative).

Statistical analyses were performed in Stata/MP, version 15.2 (StataCorp, CollegeStation, TX) and R statistical software.

## RESULTS

Of the 214 specimens from potential CCP donors, 146 were tested by the Euroimmun, EDI, and Abbott assays; 140 were tested by the ImmunoDiagnostics assay, and all 214 were tested by the Roche assay (140 were assayed by all five EIAs). Of the 1,102 pre-pandemic specimens included, 562 were tested by the Euroimmun assay, 579 were tested by the EDI assay, 306 were tested by the ImmunoDiagnostics assay, and 500 were tested by the Abbott and Roche assays. In empirical ROC analyses, all assays —with the exception of EDI— had an AUROC value that exceeded 0.95, suggesting each assay has the capacity to accurately detect antibodies to SARS-CoV-2 (**Table 2**). For the ELISAs (Euroimmun, EDI, and ImmunoDiagnostics) the AUROCs were greater by 5 absolute percentage points in the empirical ROC analysis compared to the analysis using the manufacturers’ cutoffs. For the Abbott and Roche assays, the AUROCs were similar in the empirical analysis and the analysis using the manufacturers’ cut-offs.

**Table 2.**
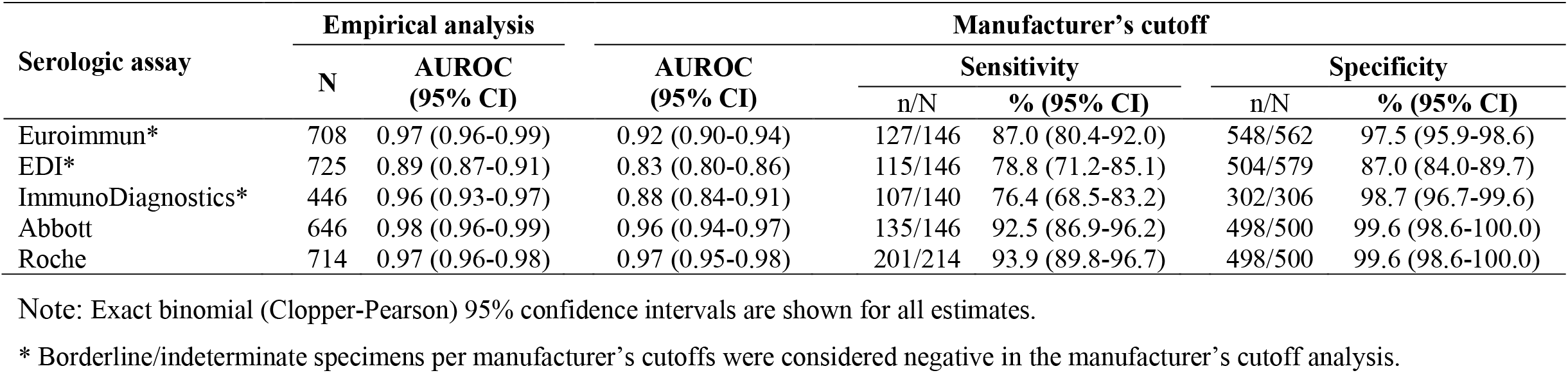
Diagnostic accuracy of various enzyme immunoassays to detect IgG or total antibodies to SARS-CoV-2.

Using the manufacturers’ cut-offs, the sensitivity of each EIA to detect SARS-CoV-2 antibodies ranged from 76.4% to 93.9%, whereas the specificity of each EIA ranged from 87.0% to 99.6%. Both the Abbott and Roche assays had comparable characteristics as each other with higher point estimates for sensitivity and specificity compared to the ELISAs. Considering indeterminate/borderline specimens as seropositive as opposed to seronegative decreased the specificity of EDI; however, excluding indeterminate/borderline specimens had minimal impact on estimates ***(Supplemental Table 2)***. Similar estimates were also obtained in direct comparisons ***(Supplemental Tables 3***, 4). It is also notable that among the 140 CCP donor specimens that were tested by all five EIAs, there were 6 (4.3%) specimens that were seronegative (or indeterminate) for SARS-CoV-2 by all five EIAs. The median time from COVID-19 diagnosis for these 6 individuals was 46 days (range, 33-54). Interestingly, there were 2 false-positive specimens of the 500 pre-pandemic specimens tested by both Abbott and Roche (one of which was false-positive on both assays).

Among pre-pandemic samples, there was greater variation in the distribution of ratio values for ELISAs than for the Abbott and Roche assays ***(Supplemental Figure*** 1), consistent with the higher specificity observed for the Abbott and Roche assays. For the Abbott, Roche and ImmunoDiagnostics assays, the value of three times the standard deviation above the mean value from all the pre-pandemic samples was below the cutoff used to define a positive sample.

Among the 140 CCP donor specimens, the median nAb AUC value was 60 (interquartile range: 10, 150). The prevalence of nAb AUC ≥20 was 65.7% (n=92), the prevalence of nAb AUC ≥40 was 57.1% (n=80), the prevalence of nAb AUC ≥80 was 45.7% (n=64), and the prevalence of nAb AUC ≥160 was 25.0% (n=35). There were significant positive correlations between nAb AUC values and EIA ratio values for all EIAs examined (**Figure 1**), but the strongest correlation was observed for the Euroimmun assay (*ρ*=0.81 [95%CI: 0.74-0.85]) and weakest correlation was observed for the Roche assay (*ρ*=0.40 [95%CI: 0.25-0.54]). With “high” nAb titers as the reference positive, there was substantial between-assay variability in the empirical AUROCs of each EIA, but changing the threshold used to define a “high” nAb titer did not substantially impact the AUROCs of a given EIA (**Figure 2**). For instance, for all four nAbs thresholds evaluated, all empirical AUROC point estimates for the Euroimmun assay were ≥90, whereas all AUROC point estimates for the Roche assay were <0.75. For the Euroimmun assay and nAB test at a threshold of ≥160, the EIA ratio cut-off with the highest overall percent agreement (86%) was 6.0 (positive percent agreement was 77% and negative percent agreement was 89%).

**Figure 1.**
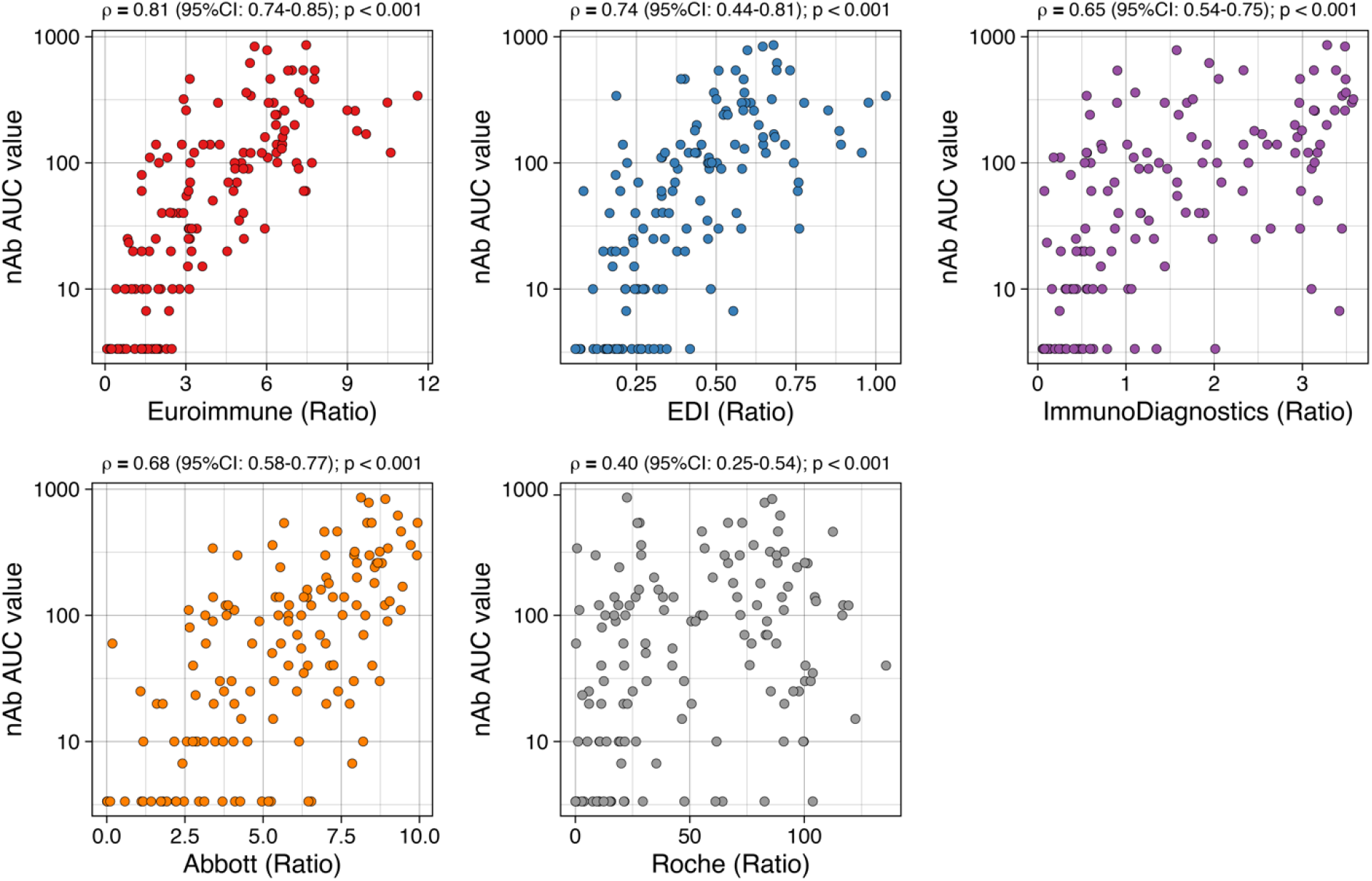
Correlations between SARS-CoV-2 enzyme immunoassay antibody titers and neutralizing antibody titer AUC values in COVID-19 convalescent individuals (n=140). Spearman correlation coefficients (*ρ*) were calculated with 95% confidence intervals (CI) estimated over 1000 bootstrap iterations.

**Figure 2.**
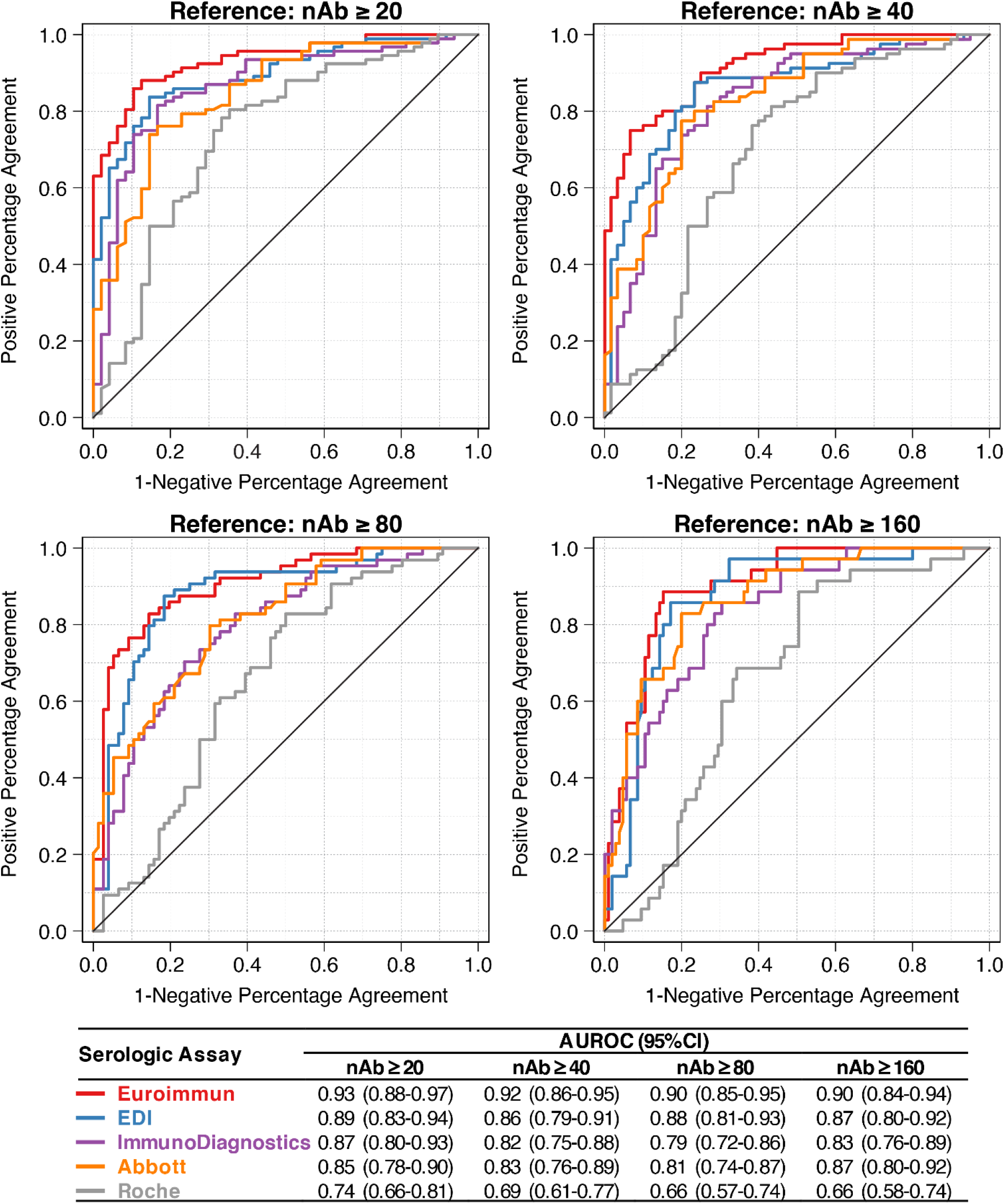
Empirical receiver operating curve analysis for various SARS-CoV-2 enzyme immunoassays to detect high neutralizing antibody (nAb) titers at various thresholds (n=140). Four thresholds for a high nAb AUC value were examined as the reference positive test.

**Table 3** shows the positive percentage agreement (sensitivity) and negative percentage agreement (specificity) of each assay with the four nAb test thresholds when using the EIA manufacturers cut-offs for seropositivity. All EIAs had a positive percent agreement with “high” nAbs exceeding 90%, regardless of the threshold for high nAbs. However, there was poor negative percentage agreement between each EIA and nAbs. For all EIAs, the negative percentage agreement decreased with increasing threshold for high nAbs.

**Table 3.** Concordance between manufacturer enzyme immunoassay cut-offs for SARS-CoV-2 seropositivity and high neutralizing antibody titers at various thresholds.

| **Serologic assay** | **Positive Percentage Agreement, no. (%)** |  |  |  |
| --- | --- | --- | --- | --- |
|  | **nAb ≥20**<br>(n=92) | **nAb ≥40**<br>(n=80) | **nAb ≥80**<br>(n=64) | **nAb ≥160**<br>(n=35) |
| Euroimmun | 90 (97.8%) | 80 (100%) | 64 (100%) | 35 (100%) |
| EDI | 86 (93.5%) | 74 (92.5%) | 61 (95.3%) | 34 (97.1%) |
| ImmunoDiagnostics | 86 (93.5%) | 76 (95.0%) | 61 (95.3%) | 35 (100%) |
| Abbott | 90 (97.8%) | 79 (98.8%) | 64 (100%) | 35 (100%) |
| Roche | 90 (98.4%) | 78 (97.5%) | 63 (98.4%) | 34 (97.4%) |
| **Serologic assay** | **Negative Percentage Agreement, no. (%)** |  |  |  |
|  | **nAb <20**<br>(n=48) | **nAb <40**<br>(n=60) | **nAb <80**<br>(n=76) | **nAb <160**<br>(n=105) |
| Euroimmun | 16 (33.3%) | 18 (30.0%) | 18 (23.7%) | 18 (17.1%) |
| EDI | 25 (52.1%) | 25 (41.7%) | 28 (36.8%) | 30 (28.6%) |
| ImmunoDiagnostics | 27 (56.3%) | 29 (48.3%) | 30 (39.5%) | 33 (31.4%) |
| Abbott | 9 (18.8%) | 10 (16.7%) | 11 (14.5%) | 11 (10.5%) |
| Roche | 6 (12.5%) | 6 (10.0%) | 7 (9.2%) | 7 (6.7%) |

## DISCUSSION

We observed substantial variability in the performance characteristics of five commercially available EIAs for the detection of antibodies to SARS-CoV-2 and detection of high nAb titers in convalescent individuals. The Roche and Abbott assays had high diagnostic accuracy for the detection of antibodies against SARS-CoV-2. However, the Roche assay ratios weakly correlated with nAb titers and poorly identified persons with high nAb titers. In contrast, the Euroimmun assay ratios had the highest correlations with nAb titers and high discriminative capacity for detecting high nAbs. This variability in assay performance should be considered when selecting an EIA to detect antibodies against SARS-CoV-2 and/or high nAbs among recovered persons.

Consistent with our findings, there is growing evidence that both the Abbott and Roche assays have comparable performance characteristics that are often superior to many other commercially available ELISAs to detect IgG or total antibodies against SARS-CoV-2 in convalescent individuals.^19,20^ Although we did not include “challenge” specimens to examine potential crossreactivity of antibodies to other pathogens, others have shown limited evidence of crossreactivity for the Euroimmun, EDI, Roche, and Abbott assays.^19,21-25^ Data on the performance of the ImmunoDiagnostics ELISA to detect SARS-CoV-2 antibodies are limited.

Large public health laboratories and large blood collection centers often rely on automated serological platforms—like those by Roche and Abbott—for screening of multiple pathogens including SARS-CoV-2. While our data support the use of Roche and Abbott to detect SARS-CoV-2 antibodies, their utility to detect high nAbs in CCP donors is less clear. Similar to prior reports, we observed varying degrees of positive correlations between commercial EIA ratios and neutralizing titers.^14,26^ It is perhaps unsurprising that the Euroimmun ELISA ratios correlated best with nAb titers since it detects S1-specific antibodies—a subset of which are responsible for virus neutralization—while the other assays we assessed detect N-specific antibodies which lack virus neutralization activity. Accordingly, our empirical ROC analysis also indicates Euroimmun may have better performance in discriminating high nAb titers, as compared to the Abbott and Roche assays. Interestingly, using the manufacturer’s cut-off, Jaaskelainen et al. found the Abbott assay had greater positive and negative percent agreement with nAb activity than the Euroimmun assay.^27^ In our study, the Abbott assay was also better able to discriminate high nAbs than the Roche assay, which is in contrast to a study by Tang et al. that found similar performance between the Abbott and Roche assays.^28^ However, similar to Tang et al., we found that applying the manufacturer’s cutoffs for the commercial EIAs (including Euroimmun) led to suboptimal negative percentage agreement with high nAbs near the FDA recommended nAb titer cut-off of ≥1:160.^28^ Larger comparative studies are needed to determine the optimal EIA and cut-off to discriminate nAb levels in convalescent donors, including other promising EIAs that were not included in these evaluations.^29^

This study has limitations. First, the data were cross-sectional, so we were unable to capture the influence of longitudinal antibody dynamics on diagnostic accuracy. Second, there were several types of specimens that were not included in the evaluation, such as samples from early in SARS-CoV-2 infection (e.g., <14 days post-symptom onset), samples from individuals who were asymptomatic when infected with SARS-CoV-2, and samples from convalescent individuals who were infected >6 months ago—all of which could potentially influence our estimates of assay sensitivity. Third, the samples used to examine assay specificity were not well-characterized due to the identity-unlinked design of the JHHED serosurvey. However, given that we used samples from patients in an inner-city emergency department that delivers primary care to the local underserved community, several included patients who were likely seeking care for viral respiratory illnesses. Finally, the samples evaluated were primarily from the Baltimore-Washington D.C. region, and results may not be generalizable elsewhere.

Implementation of the appropriate EIAs to detect SARS-CoV-2 antibodies will require careful consideration of the inferential purpose (e.g., individual-vs. population-level inference), context (e.g., prevalence in target population), operational feasibility (e.g., high-throughput platform vs. manual ELISA) and the underlying test performance characteristics of the assays. Although the output ratio results for commercially available EIAs correlate with nAb titers, EIA ratios should not be universally considered a surrogate for nAb titers. This is particularly relevant for programs that are currently scaling CCP therapy per new FDA guidelines. Ratios from some commercial EIAs, however, may help inform prediction models that can also incorporate other predictors of high nAb titers. These models could prove useful in the identification of optimal CCP donors in the absence of accurate and reliable high-throughput tests for nAb titers.

## Data Availability

Data is available upon request

## Acknowledgements

We acknowledge all of the participants who contributed specimens to this study and all study staff without whom this study would not have been possible.

## Funding

This work was supported in part by the Division of Intramural Research, National Institute of Allergy and Infectious Diseases (NIAID), National Institutes of Health, as well as extramural support from NIAID (R01AI120938, R01AI120938S1 and R01AI128779 to **A.A.R.T**; 298 R01AI05273 and R01AI152078 to **A.C**.; T32AI102623 for supporting **E.U.P**.; UM1‐AI068613 for supporting **R.E.F**. and **E.K**.; and NIH Center of Excellence in Influenza Research and 300 Surveillance HHSN272201400007C to **A.P**. and **R.E.R**.), National Heart Lung and Blood Institute (K23HL151826 to **E.M.B** and R01HL059842 to **A.C**.), National Institute of Drug Abuse (T32DA007292 for supporting **D.B**.), Bloomberg Philanthropies (**A.C**.) and Department of Defense (W911QY2090012 to **A.C**. and **D.S**.).

## Conflicts of interest

The authors declare no potential conflicts of interest.

## Notes

### Competing Interest Statement

The authors have declared no competing interest.

### Author Declarations

This study used stored samples and data from two parent studies that were approved by The Johns Hopkins University School of Medicine Institutional Review Board.

